# Heterozygous and homozygous variants in *STX1A* cause a neurodevelopmental disorder with or without epilepsy

**DOI:** 10.1101/2022.04.20.22274073

**Authors:** Johannes Luppe, Heinrich Sticht, François Lecoquierre, Alice Goldenberg, Kathleen Gorman, Ben Molloy, Emanuele Agolini, Antonio Novelli, Silvana Briuglia, Outi Kuismin, Carlo Marcelis, Antonio Vitobello, Anne-Sophie Denommé-Pichon, Sophie Julia, Johannes R. Lemke, Rami Abou Jamra, Konrad Platzer

**Affiliations:** Institute of Human Genetics, University of Leipzig Medical Center, Leipzig, Germany; Institute of Biochemistry, Friedrich-Alexander-Universität Erlangen-Nürnberg, Erlangen, Germany; Department of Genetics and Reference Center for Developmental Disorders, Normandie Univ, UNIROUEN, CHU Rouen, Inserm U1245, FHU G4 Génomique, F-76000, Rouen, France; Department of Neurology and Clinical Neurophysiology, Children’s Health Ireland at Temple Street, Dublin; School of Medicine and Medical Science, University College Dublin, Dublin, Ireland; Genuity Science, Dublin, Ireland; Laboratory of Medical Genetics, Bambino Gesù Children Hospital IRCCS, Rome, Italy; Department of Biomedical, Dental, Morphological and Functional Imaging Sciences, University of Messina, Italy; Institute for Molecular Medicine Finland, Helsinki, Finland; Department of Human Genetics, Radboud University Medical Center, Nijmegen, Netherlands; University of Burgundy-Franche Comté, Dijon, France; Federative Institute of Biology, CHU de Toulouse, Toulouse, France; Center for Rare Diseases, University of Leipzig Medical Center, Leipzig, Germany

**Keywords:** Epileptic encephalopathy, SNAREopathy, Synapse

## Abstract

The neuronal SNARE complex drive synaptic vesicle exocytosis. Therefore, one of its core proteins syntaxin 1A (STX1A) has long been suspected to play a role in neurodevelopmental disorders. We assembled eight individuals harboring rare variants in *STX1A* who present with a spectrum of intellectual, autism and epilepsy. Causative variants comprise a homozygous splice variant, three *de novo* missense variants and two inframe deletions of a single amino acid. We observed a phenotype mainly driven by epilepsy in the individuals with missense variants in contrast to intellectual disability and autistic behavior in individuals with single amino acid deletions and the splicing variant. *In silico* modeling of missense variants and single amino acid deletions show different impaired protein-protein interactions. We hypothesize the two phenotypic courses of affected individuals to be dependent on two different pathogenic mechanisms: (1) a weakened inhibitory STX1A-STXBP1 interaction due to missense variants results in an *STX1A*-related developmental epileptic encephalopathy and (2) a hampered SNARE complex formation due to inframe deletions causes an *STX1A*-related intellectual disability and autism phenotype.

## Introduction

SNARE (soluble NSF attachment protein receptor) complexes play a crucial role in a multitude of membrane fusion processes in the central nervous system. The core complex, comprising neuronal SNAREs syntaxin 1, SNAP25 and VAMP2, mediates synaptic vesicle exocytosis^1^ as well as secretion of neuropeptides and neurotrophins.^2^ Syntaxin 1A (encoded by the gene *STX1A*) is localized at the neuronal synaptic plasma membrane. It is involved in regulation of several plasma membrane-bound monoamine transporters.^3–6^ Variants in *STX1A* have not been shown to cause a mendelian disorder. Of note, *de novo* deletions of *STX1A* was seen in 5 out of 83 individuals with autism spectrum disorder (ASD)^7^ although a recent analysis of a large ASD cohort did not show a significant association.^8^ Single nucleotide polymorphisms in *STX1A* are suspected to increase susceptibility to migraine in several case-control-studies^9–12^ as well as playing a role in predisposition to autism^13–15^, cryptogenic epilepsy^16^ and children attention-deficit/hyperactivity disorder^17^, whereas no associations could be drawn to schizophrenia.^18^ Due to little redundancy and rescue mechanisms, disruption of components of the SNARE complex give rise to a group of rare diseases called SNAREopathies.^19,20^ In this study, we describe eight individuals harboring rare homozygous or *de novo* heterozygous variants in *STX1A* to delineate a novel *STX1A*-related neurodevelopmental disorder based on different pathogenic mechanisms and inheritance models.

## Materials (Subjects) and Methods

### Standard protocol approvals

The study was approved by the ethics committee of the University of Leipzig, Germany (402/16-ek). All families provided informed consent for clinical phenotyping, genetic testing and publication. If done in a research setting, testing was approved by local ethics committees in the respective institutions.

### Research cohort and identification of variants

Eight individuals with *STX1A* variants from seven different families were assembled via literature review and GeneMatcher.^21^ Phenotypic and genotypic information was obtained from the referring collaborators using a standardized questionnaire, evaluating family anamnesis, clinical history, genetic testing, variant details, EEG, brain imaging and medication. All individuals underwent exome or genome sequencing. Since no causative variants in a known disease gene were identified, the data were examined in a scientific approach, including parental sequence data if available. All variants were prioritized considering allele frequency in gnomAD^22^ below 1%, impact in different *in silico* programs and involvement of candidate genes in neuronal processes. All variants in *STX1A* are described with regard to GRCh37 and NM_004603.3.

### Structural modeling

Structural analysis of the variants was performed based on the crystal structure of Syntaxin 1A in complex with Syntaxin-binding protein 1 (PDB: 3C98)^23^ or the components of the SNARE complex (PDB: 6IP1, 6MDN).^24,25^ Amino acid changes were modeled with SwissModel^26^ and RasMol^27^ was used for structure analysis and visualization.

## Results

### Clinical description

The eight individuals exhibited varying neurodevelopmental, neurological and neuropsychiatric symptoms. In four out of eight individuals, epilepsy was the leading clinical symptom, whereas in the four remaining individuals intellectual disability and autistic behavior without epilepsy was prominent. An overview of the clinical symptoms is presented in Table 1 and Table S1.

**Table 1:** Summary of clinical symptoms of individuals with rare variants in STX1A

| Ind | variant | origin | sex | ID | motor delay | epilepsy | neurological symptoms, regression | behaviour | other symptoms |
| --- | --- | --- | --- | --- | --- | --- | --- | --- | --- |
| 1 | c.284-1G>A, homozygous | parental | m | moderate | yes | no | neonatal hypotonia, no regression | no ASD, behavior unremarkable | unremarkable |
| 2 | c.284-1G>A, homozygous | parental | m | moderate | yes | no | neonatal hypotonia, no regression | no ASD, aggressiveness | unremarkable |
| 3 | p.(Cys145Trp), heterozygous | de novo | m | severe | yes | yes, epileptic encephalopathy, generalized tonic-clonic seizures | hypotonia, ataxia, spasticity, pyramidal signs, speech regression | ASD, stereotypes, drooling, bruxism, likes pulling the hair, biting | ptosis |
| 4 | p.(Cys145Trp), heterozygous | de novo | f | severe | yes | yes, absence seizure, generalized tonic-clonic | sensory processing disorder, regression | features of ASD, but did not fulfill criteria | Tight ankles, diazoxide responsive congenital hyperinsulinism |
| 5 | p.(Ser185Cys), heterozygous | de novo | m | moderate | yes | yes, rolandic epilepsy | neonatal hypotonia, fine tremor, no regression | no ASD, aggressiveness and hyperactivity | congenital clubfoot, brachycephaly, ears low set, retrognathia, tapered fingers, lower limb muscular hypotrophy and hypotenar eminence, hypochromic and coffee-milk skin spots. |
| 6 | p.(Val223del), heterozygous | unknown |  | moderate |  | no | NA | NA | NA |
| 7 | p.(Gln226Arg), heterozygous | de novo | f | severe | yes | yes, West syndrome, Lennox-Gastaut syndrome | neonatal hypotonia, dysphagia, spastic paresis | unremarkable | drooling |
| 8 | p.(Val241del), heterozygous | de novo | m | severe | yes | no | neonatal hypotonia, no regression | ASD, extreme aggressiveness | pes varus, constipation |

ASD: autism spectrum disorder; ID: intellectual disability; NA: not available

#### Individual 1

This male was first reported in Reuter et al.^28^ He was born to consangineous parents with normal intelligence. One of his brothers (described below as individual 2) presented with similar symptoms. Another brother had died in infancy (see Figure S1). During pregnancy of individual 1 decreased fetal movements were noted. He was born at term with increased birth length and neonatal hypotonia. He learned unassisted walking at the age of and did not show persistent motor problems in later childhood. Global development and speech were delayed. Clinical assessment at the age of showed moderate intellectual disability with mostly unremarkable speech and behavior. Seizures were denied. MRI or EEG were not performed. No facial dysmorphisms were seen. Most of his time, he worked on the acre of the family.

#### Individual 2

This male was also first reported in Reuter et al.^28^ and is the younger brother of individual 1. He was born as the in the family described above. As in his brother, decreased fetal movements were noted in pregnancy. After birth at term, neonatal hypotonia and increased birth length were noted. He also learned unassisted walking around age and did not show persistent motor problems in later childhood. Global development and speech were delayed. Clinical assessment at the age of showed moderate intellectual disability, but a mostly unremarkable speech. His behavior was more aggressive compared to his brother. No seizures were recorded. Neither MRI nor EEG were performed. No facial dysmorphisms were seen. As his brother he mostly worked on the acre of the family.

Exome sequencing detected a homozygous variant in *STX1A*:c.284-1G>A in both brothers. The parents and one of the sisters were found to be heterozygous for this variant via Sanger sequencing. Unfortunately, this family was lost to further follow-ups.

#### Individual 3

This male was born after an unremarkable pregnancy to healthy parents. Birth measurements were around the 50th percentile and neonatal hypotonia was observed. This individual was affected by severe global developmental delay. He learned unassisted walking at the age of, but had persistent motor problems in later childhood. Clinically, he was suspected to harbor an Angelman-like disorder. He was able to speak two to three words at the age, but speech was absent at last clinical assessment at the age of . His intellectual disability was assessed to be severe. Abnormal behavior comprised stereotypes, drooling, bruxism, biting and pulling hair and he was diagnosed with ASD. He also had recurrent sleeping problems. Neurological examination showed ptosis, hypotonia, ataxia and spasticity with pyramidal signs. First tonic-clonic seizures were noticed at the age of, reaching a frequency of several per day. A therapy with valproate and levetiracetam was successful to suppress severe seizures for several years, although absences and rare tonic seizures remained. At age of seizures intensified again, reaching frequencies of several tonic and tonic-clonic seizures and absences per day. A switch in medication to rufinamide, valproate and perampanel lead to a limited response with suppressing seizure frequency to one per week. MRIs were normal apart from delayed myelination at three years of age. Family history only elicited a febrile seizure in his maternal uncle at the age of.

Exome sequencing detected a heterozygous *de novo* variant in *STX1A*:c.435C>G, p.(Cys145Trp).

#### Individual 4

A girl, was born after an uneventful pregnancy at 39 weeks of gestation. Birth weight was on the 75^th^ percentile. First seizures occurred on, which was attributed to neonatal hypoglycemia (1.4mmol/L glucose) due to diazoxide responsive congenital hyperinsulinism. She was in the neonatal intensive care unit. After the neonatal period, she was seizure-free until of age, when she presented with febrile seizures, and shortly after recurrent afebrile focal seizures (2-3 per week). Subsequently she developed focal to bilateral tonic-clonic, absence and generalized onset motor tonic-clonic seizures. At age of, she presented with myoclonic, myoclonic absence and focal onset (occipital lobes) seizures. Pharmacological treatment included valproate, phenobarbitone, pyridoxine, levetiracetam, carbamazepine, lamotrigine, zonisamide, clobazam, lacosamide, rufinamide, ethosuximide. Her current seizure frequency is approximately 2 absences and 1 focal seizure per month on sodium valproate and ethosuximide. Age of, weight is on the 50^th^ to 75^th^ percentile. Concerning her cognitive development she started babbling aged but then stopped at indicating a regression of skills. She has severe ID and never acquired speech, but can use picture books to communicate. She walked independently aged, but has persistent motor problems in later childhood with a waddling gait. She has autistic traits but does not meet the criteria for an ASD diagnosis and she was diagnosed with sensory processing disorder. The latest EEG at of age showed a slow background, multifocal epileptiform spikes, and occipital spike waves and generalized irregular-semi-irregular spike waves. MRI performed at of age showed mild periventricular leukoencephalopathy. In family history, the mother had two seizures as a teenager and was treated with Carbamazepine for two years, after which she was weaned. No further seizures occurred.

Trio genome sequencing detected a heterozygous *de novo* variant in *STX1A*: c.435C>G, p.(Cys145Trp).

#### Individual 5

This male was born at term after an unremarkable pregnancy. His birth length was 49 cm (14^th^ percentile), weight 2800 gram (12^th^ percentile) and head circumference 34 cm (17th percentile). Neonatal hypotonia was noticed. He had global developmental delay and persistent motor problems throughout his life, but learnt unassisted walking at the age of. His first speech was noted at the age of. At the age of, his speech was mildly age-inappropriate, reaching 5^th^ centile (Bayley Scales of Infant Development). Moderate intellectual disability as well as behavioral problems such as aggressiveness, hyperactivity, attention deficit and sleeping problems were reported. He had first seizures (loss of consciousness) at the age of with a frequency of one seizure every two to three months. He exhibited right frontal to centero-temporal spikes in EEG and was diagnosed with rolandic epilepsy. Antiepileptic therapy was started at the age of and consisted of Levetiracetam. During therapy, no further seizures occurred, but the EEG anomalies persisted. Neurological examination showed fine intentional tremors as well as global motor impediment. A brain MRI at the age of had normal results. Furthermore, axial hypotonia, a congenital clubfoot and various mild dysmorphisms, including brachycephaly, low set ears, retrognathia, tapered fingers, lower limb muscular hypotrophy and hypotenar eminence as well as hypochromic and coffee-milk skin spots were noted. His weight reached 97th percentile, OFC was at the 36th percentile. In family history, the mother was diagnosed with migraine and a Chiari malformation.

Trio exome sequencing detected a heterozygous *de novo* variant in *STX1A*: c.554C>G, p.(Ser185Cys).

#### Individual 6

This patient had moderate intellectual disability. No seizures were recorded. Unfortunately, this patient was lost to follow-up for further detailed phenotyping. Exome sequencing detected a heterozygous variant in *STX1A*: c.668_670delTGG. p.(Val223del) of unknown origin due to unavailable parental testing.

#### Individual 7

This female was born to healthy parents after 42 weeks of an unremarkable pregnancy. Birth weight was 3300 g, neonatal hypotonia was noted. At the age of, body measurements were: 6085 g (11^th^ percentile), 66 cm (46^th^ percentile) and OFC 41cm (9^th^ percentile). At last follow-up at the age of measurements were: 38 kg (2nd percentile), 150 cm (3rd percentile). She had severe global developmental delay, motor problems (she never learned walking), absence of speech and profound intellectual disability. Neurological examination showed dysphagia, drooling and spastic paresis.

Epileptic spasms became evident at the age of and were associated with hypsarrhythmia on EEG, compatible with West syndrome. Additionally, daily tonic seizures occurred and a severe epileptic encephalopathy developed. Treatment included ACTH, valproate, levetiracetam, clonazepam and cannabidiol, suppressing frequency to weekly intervals but without reaching longer seizure free periods. The encephalopathic phenotype later progressed to Lennox-Gastaut syndrome. A brain MRI at the age of three years showed normal results.

Trio exome sequencing detected a heterozygous *de novo* variant in *STX1A*: c.677A>G, p.(Gln226Arg).

#### Individual 8

This male was born at term to healthy parents after an unremarkable pregnancy. Birth measurements determined a length of 50 cm (8^th^ percentile) and a weight of 3000 g (14^th^ percentile). Neonatal hypotonia was observed. At age of he showed a length of 71 cm (< 1st percentile) and a weight of 9 kg (1st percentile). At of age length was at -3 SD while OFC was at +2 SD. This individual was affected by severe intellectual disability with only few words at last assessment and motor delay with unassisted walking at around four years of age. Behavioral problems compatible with an autism spectrum disorder were noted, including an extreme aggressiveness and sleeping problems. Apart from neonatal hypotonia no further neurological abnormalities could be elicited. The individual had no seizures and brain MRI showed normal results. Additional symptoms included a pes varus and constipation.

Trio exome sequencing showed a heterozygous *de novo* variant in *STX1A*: c.722_724delTAG, p.(Val241del).

### Genotypic spectrum

One homozygous splicing variant, three different *de novo* missense variants and two deletions of a single amino acid (one *de novo*) were observed in *STX1A*. Variants were predicted to be damaging according to multiple *in silico* tools (Tables S2 and S3). The only recurrent variant is the missense variant p.(Cys145Trp) twice occurring *de novo* in individuals with a consistent phenotype.

Furthermore, in the most recent DDD study^29^, the *de novo* variant c.236T>G, p.(Met79Arg) was reported in an individual with a neurodevelopmental disorder, but a detailed phenotypic description is not available.

### Structural modeling

All residues affected by a variant in the present study are resolved in the crystal structure of STX1A in complex with syntaxin-binding protein 1 (STXBP1).^23^ In this complex, residues 29-237 of STX1A form a rather compact structure comprising four helices (Fig. 2A). This conformation was termed ‘closed’ conformation in the literature.^23^ The major effect of the missense variants are steric clashes, which destabilize the STX1A structure and are therefore expected to hamper interaction with STXBP1. A similar negative effect is expected for the deletion of p.(Val223del), which disrupts the helical structure of the H3 domain (residues 186-253). In contrast, p.(Val241del) plays only a minor role for binding to STXBP1. An experimental investigation proved that deletion of syntaxin1A residues 241-262 caused only a very moderate decrease in STXBP1 affinity by a factor of two^23^.

**Fig. 1:**
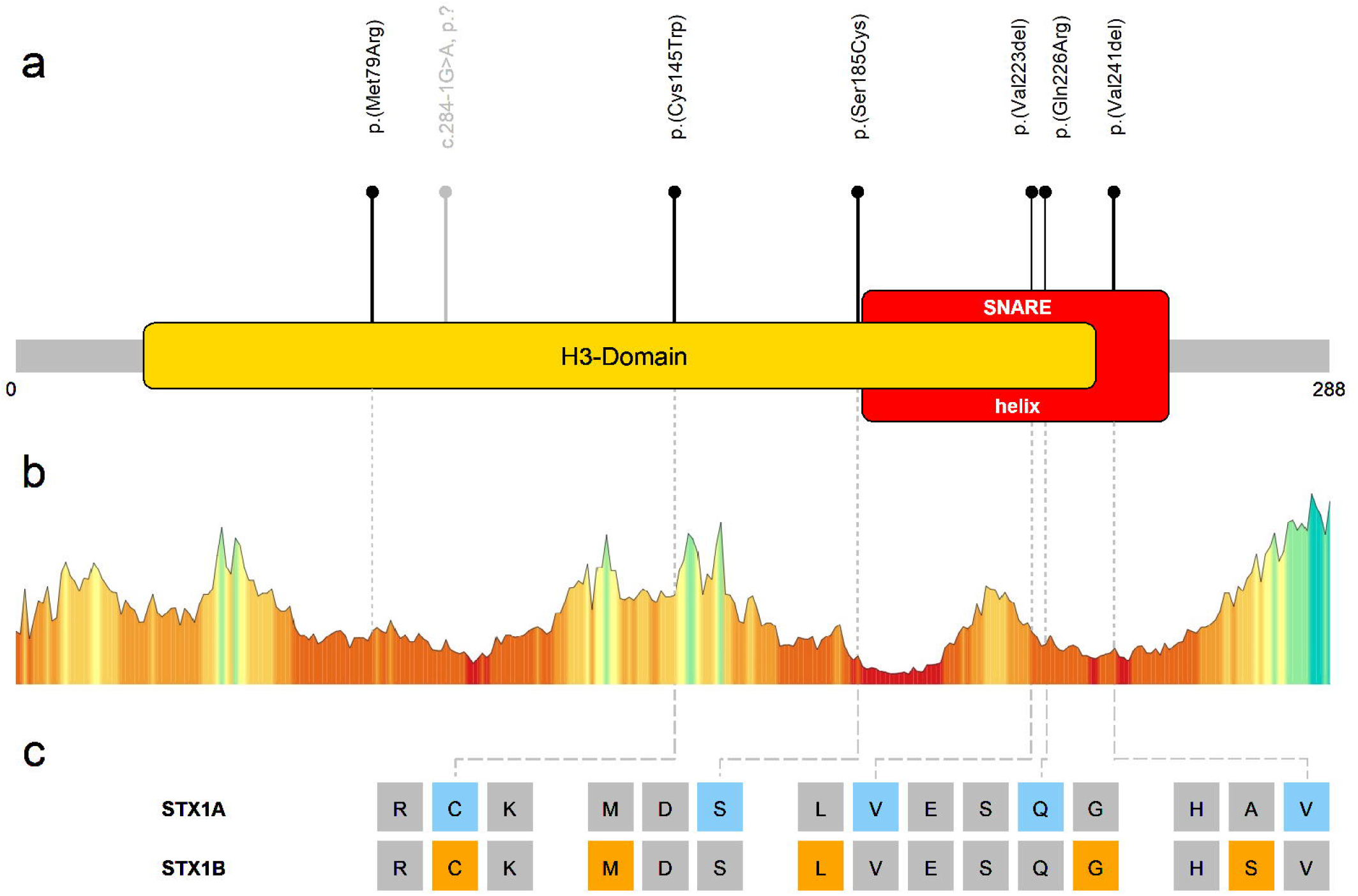
Overview of variants in *STX1A*. (a) Schematic depiction of STX1A. Segments of particular functional importance are displayed colored and named. The lollipop bars indicate observed causative variants of this study (b) Association of observed variants with a tolerance landscape of STX1A created by Metadome^42^. Height and color of the graph indicate tolerance towards variation in the referring amino acid residues: the lower and redder the bar, the less tolerant to variation is the specific residue. (c) Comparison of selected homologous amino acid residues of STX1A and STX1B. Colored positions mark residues with causative/pathogenic variants in the referring protein.

**Fig. 2:**
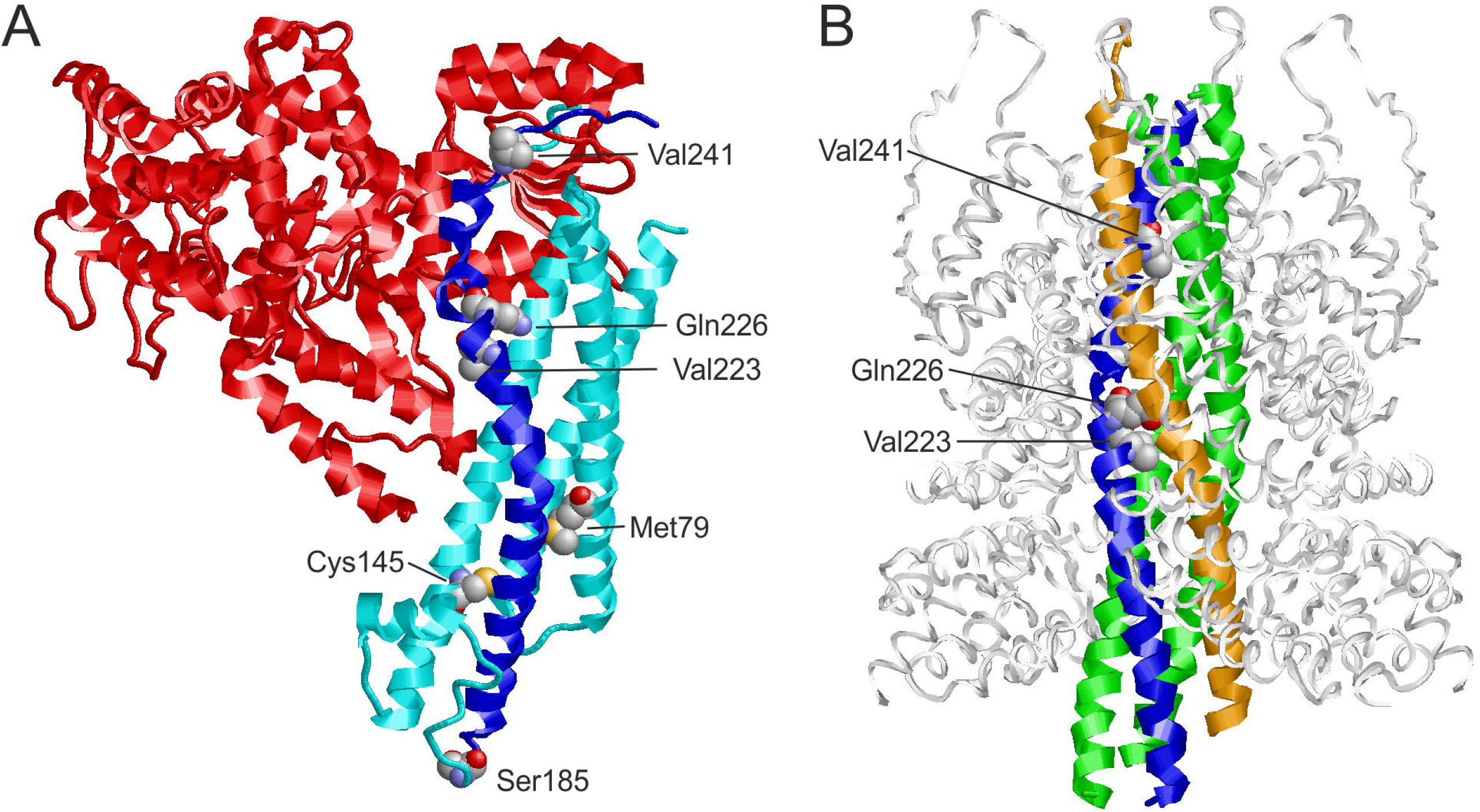
Structure of STX1A in complex with different binding partners. (a) Structure of STX1A in complex with STXBP1 (PDB: 3C98). STXBP1 and STX1A are colored in red and cyan/blue, respectively, with the H3 domain of STX1A highlighted in blue. The residues at the sites of mutation are shown in space-filled presentation and colored according to the atom types. (b) Structure of STX1A as part of the SNARE complex (PDB: 6IP1). STX1A, vesicle-associated membrane protein 2, SNAP25, and α-SNAP are colored in blue, orange, green, and white, respectively. The residues at the sites of mutation are shown in space-filled presentation and colored according to the atom types. Note that only three of the variants investigated are locate in the SNARE-interacting region.

For a structural interpretation of variant effects, it is important to note that STX1A can also adopt a structurally distinct ‘open’ conformation as part of the SNARE complex. In this complex, the H3 domain forms a long central helix that interacts with the other components of the SNARE complex (Fig. 2B). Both deletion variants p.(Val223del), p.(Val241del) are located in the central part of this helix. One-residue deletions at these sites significantly affect the helix structure and are expected to hamper SNARE complex formation. The effect of the p.(Gln226Arg) variant is known from experiment; site-directed mutagenesis revealed that this exchange results a significantly slower dissociation of the SNARE complex^30^.

## Discussion

In this study, we delineate a novel *STX1A*-related neurodevelopmental disorder based on different pathogenic mechanisms and inheritance models.

Given the data from the gnomAD database, *STX1A* is a gene with a reduced number of missense (z score = 2.4, observed/expected ratio = 0.51 (0.43 - 0.61)) and truncating/splice variants (pLI score = 0.98 observed/expected ratio = 0.06 (0.02 - 0.29)). This indicates a selective constraint on those types of variants in a healthy control population that lacks severe early onset phenotypes such as intellectual disability.^22^ All variants of this study are absent from the GnomAD database and all except the inherited splicing and the p.(Val223del) variant of unknown origin were *de novo*. Moreover, the causality of the variants is supported by strong *in silico* data^31,32^ and the structural modeling performed in this study.

In all individuals with rare variants in *STX1A*, intellectual disability, neonatal hypotonia and motor delay were present. It appears as if two different phenotypic courses are existing, following the dichotomy of the identified variants and structural modeling: (1) an epileptic encephalopathy in all individuals with missense variants and (2) a neurodevelopmental disorder with primarily intellectual disability and autistic behavior, but no epilepsy in the individuals with an inframe deletion or the family with the homozygous splice variant. The latter resembles the clinical presentation of *STX1A* haploid individuals in a Japanese ASD cohort^7^ where the probands exhibited normal or only mildly impaired intelligence with no epilepsy, but no deep phenotyping was performed. Epilepsy is also rare in individuals with Williams-Beuren-Syndrome^33^, who often lack one allele of *STX1A*, and in whom differences in transcript levels of *STX1A* have been shown to account for 15.6% of cognitive variation.^34,35^ In addition, one individual with a high quality heterozygous canonical splice site variant is listed in GnomAD_v2 and v3 respectively and therefore unlikely to be affected by intellectual disability. Thus, it appears likely that monoallelic loss of *STX1A* leads to varying defects of cognition and social behavior without severe early onset symptoms. This could explain the recessive inheritance mode in family 1 and why severe symptoms became evident in the two homozygous individuals. Epilepsy only occurs in individuals with missense variants in *STX1A* that are predicted to disturb the interaction of STX1A with STXBP1. Since both loss and gain of function variants in STXBP1 are a known cause of a developmental and epileptic encephalopathy^36^, it is tempting to speculate that the phenotypic differences in *STX1A* are also a result of loss- and gain-of-function effects in protein function. This will have to be examined in follow-up functional studies of the causative variants.

In established SNAREopathies, the most common symptoms comprise neurodevelopmental delay in domains of speech, language, motor function and intellectual ability. Additionally, other neurological symptoms such as seizures, spasms and ataxia as well as social behavioural abnormalities are observed. Severity and frequency of specific symptoms vary depending on the affected gene.^19^ The phenotype of the probands in the presented cohort in *STX1A* is therefore compatible with known SNARE-associated disorders caused by variants in *SNAP25, VAMP2, STX1B* and *STXBP1*.

Comparison of STX1A and the closely related STX1B is of special importance. The proteins show identical amino acids at more than 82 percent of positions.^37^ Mostly truncating, but also several missense variants in *STX1B* are known to cause a childhood epilepsy syndrome with both febrile and afebrile seizures, typically with a benign course and good response to treatment. Intelligence is usually normal, although more severely affected individuals have been described.^38^ Interestingly, missense variants are described to cause a more severe phenotype than null variants.^39^ For two of *STX1A* missense variants described in the present cohort, pathogenic variation in corresponding *STX1B* residues are described: In case of the *STX1A* variant p.(Cys145Trp), a missense variant affecting the homologous residue in *STX1B*: p.(Cys144Phe) was previously described as likely pathogenic.^39^ Interestingly, this individual showed additional features such as tremor and cerebellar ataxia that are not typical symptoms of *STX1B*-related generalized epilepsy with febrile seizures. For the *STX1A* variant p.(Gln226Arg) in an individual with West syndrome and Lennox-Gastaut syndrome, the homologous residue directly adjacent is affected in *STX1B*: p.(Gly226Arg) by a *de novo* variant in an individual with yet another developmental and epileptic encephalopathy with myoclonic-atonic epilepsy.^38^

Attempts to examine clinical relevance via stx1a ablated mice (null and heterozygous deletion) showed atypical social behavior and abnormal recognition profiles in a dose dependent manner.^7,40,41^ The observation of autistic symptoms in mice is another line of evidence that non-functional *STX1A*-variants rather cause a phenotype of autism and intellectual disability in humans.

STX1A adopts at least two distinct conformations; a ‘closed’ inhibitory conformation in complex with STXBP1 (Fig. 2A) and an ‘open’ conformation in the SNARE complex (Fig. 2B). In particular, the H3 domain (residues 186-253) forms entirely different interactions in both complexes. When bound to STXBP1, the H3 domain interacts with the N-terminal part of STX1A, rendering it inaccessible to its partner molecules in the SNARE complex. Therefore, incorporation of STX1A into a SNARE complex requires dissociation from STXBP1 and switch to an open conformation, in which the H3 domain is accessible.^23^ Consequently, variants may either affect the STX1A-STXBP1 complex and/or formation of the SNARE complex. Based on the structural modelling, all variants except p.(Val241del) cause a destabilization of the STX1A-STXBP1 complex thereby shifting the conformational equilibrium towards the open STX1A conformation. A reduced ability to form a functional SNARE complex is expected for the inframe deletions p.(Val223del) and p.(Val241del). In contrast, the missense variant p.(Gln226Arg) stabilizes this complex by causing a slower dissociation.^30^ These different effects on SNARE complex functioning (stabilization vs. destabilization) also offer an explanation for the different phenotype of p.(Gln226Arg) compared to the individuals harboring a deletion.

In the light of the considerations above, the phenotype of intellectual disability with epilepsy is most likely correlated with variants that weaken the inhibitory STX1A-STXBP1 interaction and do not disrupt STX1A function in SNARE complex formation. In contrast, the deletion variants, which mainly hamper SNARE complex formation, result in a different phenotype of intellectual disability and autism without seizures.

## Conclusion

In summary, different lines of evidence presented here support that rare heterozygous and homozygous variants in *STX1A* cause a neurodevelopmental disorder with two different phenotypic presentations: (1) an *STX1A*-related developmental epileptic encephalopathy due missense variants weakening the inhibitory STX1A-STXBP1 interaction and (2) an *STX1A*-related intellectual disability and autism phenotype due inframe deletions hampering SNARE complex formation. Our description thus expands the group of disorders called SNAREopathies.

## Data Availability

All data produced in the present study are available upon reasonable request to the authors.

## Abbreviations

DD/ID: developmental delay/intellectual disability
ASD: autism spectrum disorder
OFC: occipitofrontal circumference

## Acknowledgement

We thank the families for their participation and support of this study.

## Author Contributions

Conceptualization: R.A.J, K.P. Investigation: J.L, K.P.; Structural modeling: H.S.; Clinical data and genetic analysis: R.A.J., F.L., A.G., K.G., E.A., S.B., O.K., C.M., A.V., A.-S.D.-P., S.J. Visualization: H.S., J.L. Writing—original draft preparation: J.L., K.P., H.S. Review and editing of manuscript: all authors.

## Ethical Approval

The study was approved by the ethics committee of the University of Leipzig, Germany (402/16-ek).

## Supplementary material

Supplemental data are in two separate files: a .pdf-file containing Figure S1, Table S2 and Table S3. Table S1 is provided as an excel-file.

