## Supplemental Data for "Heterozygous and homozygous variants in *STX1A* cause a neurodevelopmental disorder with or without epilepsy"

**Content**

**Disclaimer:** due to medRxiv policy, all information has been removed that would allow the patient / study participant or their family, friends or neighbors to identify them (e.g., age, precise location, date of hospital admission, family relationships, past medical history, etc).

Figure S1. Pedigree of the family of Individuals 1 and 2. [removed medRxiv policy]

### Table S1. Detailed phenotypic overview of all individuals with rare variants in STX1A

Provided as a separate Excel-file. [not provided due to medRxiv policy]

### **Table S2.** *In silico* prediction of splice variant in *STX1A*

| **Ind.** | **Genomic position (hg19)** | **Variant**  **NM_004603.3** | **CADD-v6**^1^ | **SpliceAI**^2^ | **MaxEntScan**^3^ | **NNSPLICE**^4^ | **Nucleotide conservation** | **gnomAD**^5^ |
| --- | --- | --- | --- | --- | --- | --- | --- | --- |
| 1, 2 | chr7:73118754 | c.284-1G>A, p.? | 35 | 0.95 | 1 | 1 | high | 0 |

Color code represents the probability of the variant to be damaging; red: high

### **Table S3.** *In silico* prediction of missense and inframe deletion variants in *STX1A*

| **Ind.** | **Genomic position (hg19)** | **Variant**  **NM_004603.3** | **CADD-v6**^1^ | **REVEL**^6^ | **Mutation Taster**^7^ | **M-CAP 1.3**^8^ | **Polyphen 2 v2.2.2**^9^ | **AA conservation** | **gnomAD**^5^ |
| --- | --- | --- | --- | --- | --- | --- | --- | --- | --- |
| 3, 4 | chr7:73118509 | c.435C>G,  p.(Cys145Trp) | 29.2 | 0.547  (LDC) | D | 0.109  (PoP) | PrD | high | 0 |
| 5 | chr7:73117299 | c.554C>G,  p.(Ser185Cys) | 24.9 | 0.301  (MDC) | D | 0.032  (PoP) | B | high | 0 |
| 6 | chr7:73117183_73117185del | c.668_670delTGG,  p.(Val223del) | 22.1 | NA | NA | NA | NA | high | 0 |
| 7 | chr7:73117176 | c.677A>G,  p.(Gln226Arg) | 32 | 0.723  (LDC) | D | 0.328  (PoP) | PrD | high | 0 |
| 8 | chr7:73115125_73115127del | c.722_724delTAG,  p.(Val241del) | 20.4 | NA | NA | NA | NA | high | 0 |
| (55498)^11^ | chr7:73119527 | c.236T>G,  p.(Met79Arg) | 23.8 | 0.238  (MDC) | D | 0.021  (LB) | B | high | 0 |

Color code represents the probability of the variant to be damaging; red: high; orange: medium; green: low; LDC = likely disease causing; LDC = maybe disease causing; D = Deleterious; PoP = Possibly Pathogenic; PrD = Probably Damaging; B = Benign; LB = likely benign; NA = not available
